# Epidemiologial Analysis of Patients Presenting to a West London District General Hospital Requiring Admission with Covid-19

**DOI:** 10.1101/2020.10.13.20212126

**Authors:** E Heald, N Ring, D Vatvani, D Shackleton

**Affiliations:** West Middlesex University Hospital

## Abstract

**Background:** Coronavirus has lead to significant morbidity and mortality both within the UK and worldwide. We hypothesise there are local clusters of coronavirus which would therefore be amenable to targeted public health measures.

**Methods:** This is a retrospective, observational case series conducted in a West London District General Hospital. All patients admitted to hospital with a radiological or microbiological diagnosis of Covid-19 were included (children under 16 years were excluded). Consecutive sampling was used and baseline characteristics including age, sex, postcode and final patient outcome were collected from the electronic health records. Patient origin postcode was plotted to a map of the local area and an online cloud based mapping analysis system was used to generate heat maps and case density maps which were compared to living base layers. The primary outcome was identification of local clusters of cases of coronavirus. Secondary outcome was identification of population characteristics that may provide evidence for more targetted public health intervention in a second wave.

**Results:** Local clusters of infection were identified within the target population. These appeared to correlate with higher indices of deprivation, poorer overall health and high household occupancy suggesting a role for public health measures to target these areas.

**Conclusion:** There is a role for targeted public health measures in tackling the spread of coronavirus, paying particular attention to those who live in more deprived areas.

## Introduction

The novel coronavirus has dominated both scientific and mainstream news since it’s discovery in December 2020. The SARS-CoV-2 pathogen which leads to Coronavirus-19 illness (Covid-19) has resulted in more than a million deaths worldwide. There have been several studies identifying an incubation period of up to 14 days, an R value in the range of 2.6-4.71 and proven droplet, contact and aresol transmission of the virus[1]. Based on this information, governments around the world have implemented nationwide lockdown and most recently in the UK, compulsory face mask use when in an enclosed space[2]. The subsequent economic damage caused by these measures means strategies to avoid a further nationwide lockdown in the event of a second wave is a significant focus of attention. With this in mind, it is important to identify other epidemiological factors in relation to the spread of Covid-19 and those which are applicable to the UK population.

In this observational study our primary aim is to identify local clusters of patients with confirmed Covid-19 admitted to a London District General Hospital. Secondary outcomes were to identify epidemiological factors associated with local prevalence. This may provide evidence for more targeted public health interventions in the event of a second wave which in turn, will help planning of local services and resources.

## Methods

This observational study was carried out at West Middlesex University Hospital, a district general hospital on the outskirts of West London, serving a population of approximately 400,000 people. All patients presenting from 11th March to 30th April 2020 requiring admission from the Emergency Department were screened. Those that had a microbiological or radiological diagnosis of Covid-19 within 48 hours of admission were included. Paediatric patients (<16 years) were excluded. Consecutive sampling was used and baseline characteristics including age, sex, postcode and final patient outcome were collected from the electronic health record. Patients were followed up until 30th June 2020. Deaths were attributed to Covid-19 if within 28 days of a positive diagnosis and on review of clinical notes cause of death was directly linked to Covid-19. This was determined by the death certificate and when this wasn’t available, review of the clinical notes by 2 separate physicians more than 5 years qualified.

Anonymised postcode data was plotted on a map of the local area. An online cloud based mapping analysis (ArcGIS by esri) was used to generate a heat map. The same mapping analysis was used to compare case density against secondary outcome variables such as household size, indices of deprivation and perceived health.

## Results

There were 480 admissions between 11^th^ March and 30^th^ April 2020 with a diagnosis of Covid-19. Of these, 38 were excluded due to readmissions; 442 admissions were included in the analysis. There were 140 (31.7%) females and 302 (68.3%) males with an age range of 21-99 and mean age of 67. There were 182 (41.6%) deaths within the study group.

The number of admissions per day is shown in Figure 1. Admission to hospital peaked around the 1st of April, 10 days after the start of the lockdown, which is consistent with the incubation period quoted in the literature [1]. The 9th April exhibited a record 24 admissions. This aligns with the day immediately preceding a 4-day public holiday (Easter public holidays).

**Figure 1.**
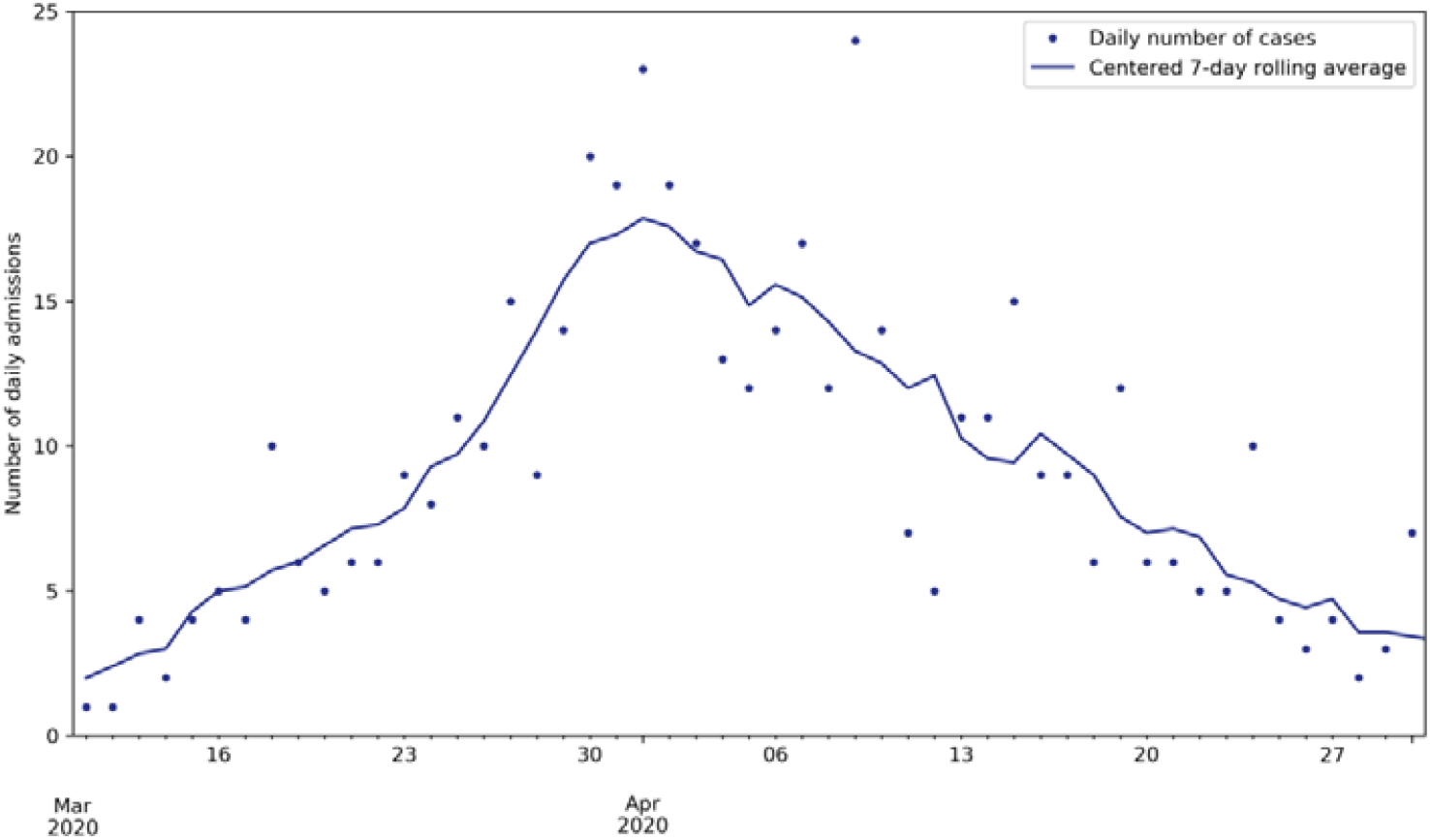
Admissions per day.

When heat mapping (Figure 2) is used to identify clusters of patients who required admission with Covid-19 we can see most areas have been affected. However, there is increased density within areas of increased conurbation, especially in the town centres of Hounslow, Isleworth, Feltham and Southall, with relative sparing around local parklands.

**Figure 2.**
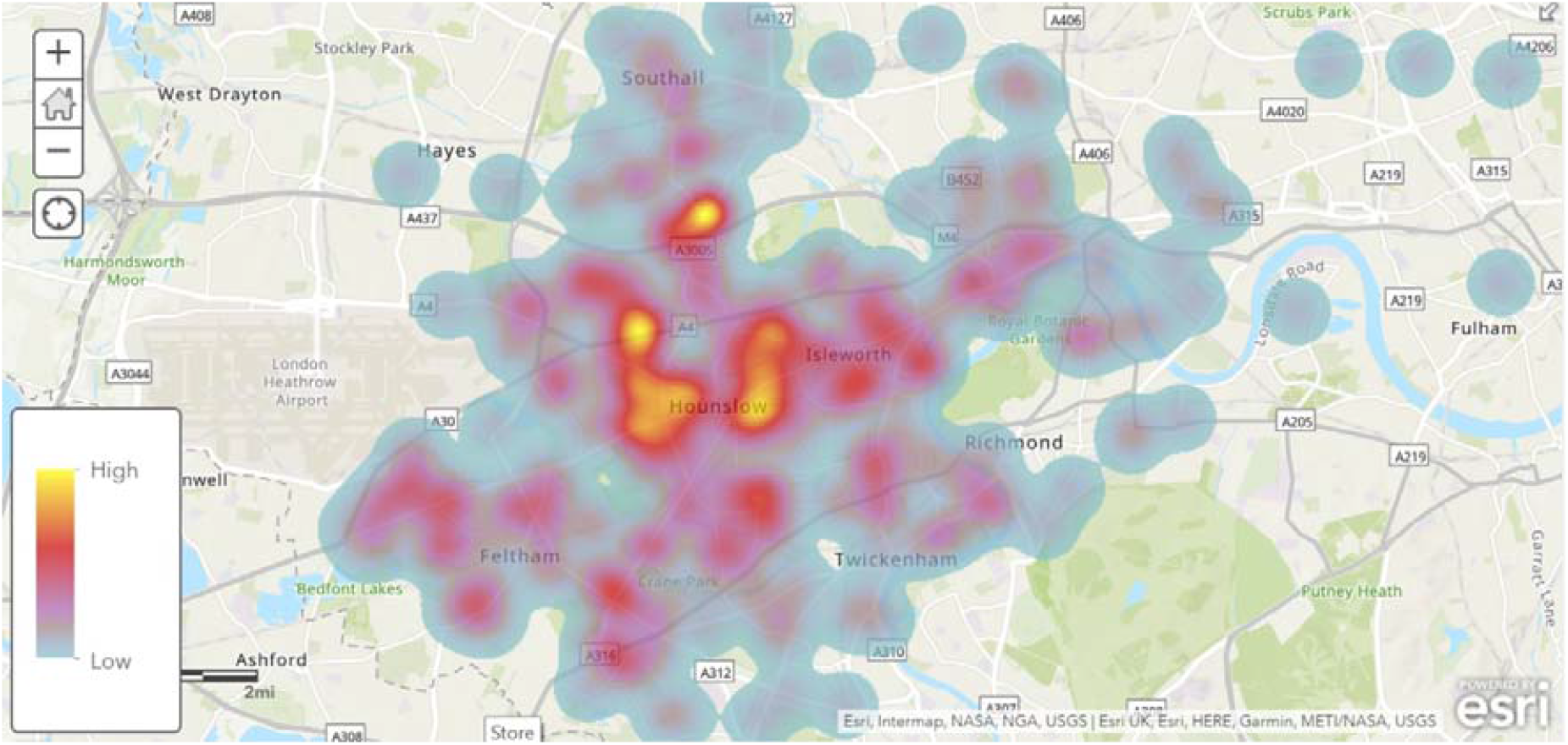
Heat Map of admissions.

When survivors and non survivors are compared (Figure 3), there is a similar spread of cases throughout the area. This suggests areas with higher admission prevalence are correlated with higher mortality, suggesting case density is a predictor of mortality.

**Figure 3.**
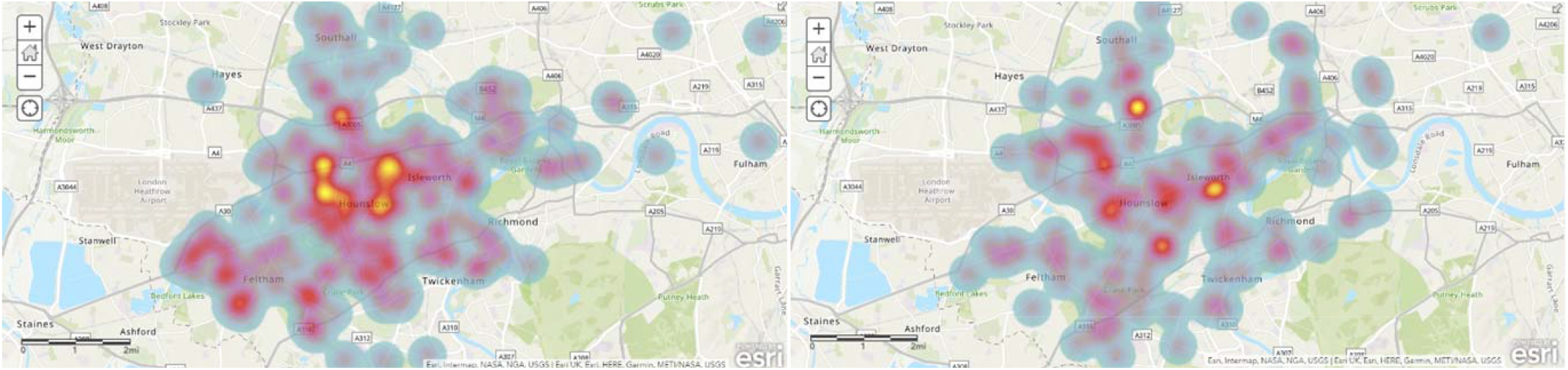
Heat Mapping of cases by survival status. Survivors (left) and non survivors (right)

When density analysis was applied, it was possible to overlay this against population maps. Index of Multiple Deprivation ranks small areas across England from 1 (most deprived) to 32844 (least deprived). In this map, this ranking has been divided into deciles with deeper reds expressing the lower deciles and therefore greatest deprivation and the deepest greens representing least deprived areas. The Index of Multiple Deprivation has been developed by The Department for Communities and Local Government and is derived from 7 domains; Income Deprivation (22.5%), Employment Deprivation (22.5%), Education, Skills and Training Deprivation (13.5%), Health Deprivation and Disability (13.5%), Crime (9.3%), Barriers to Housing and Services (9.3%), Living Environment Deprivation (9.3%) [3]. When Covid-19 admission density is mapped onto the Index of Multiple Deprivation map (Figure 4), this demonstrates maximum density does correlate with areas of greatest deprivation.

**Figure 4:**
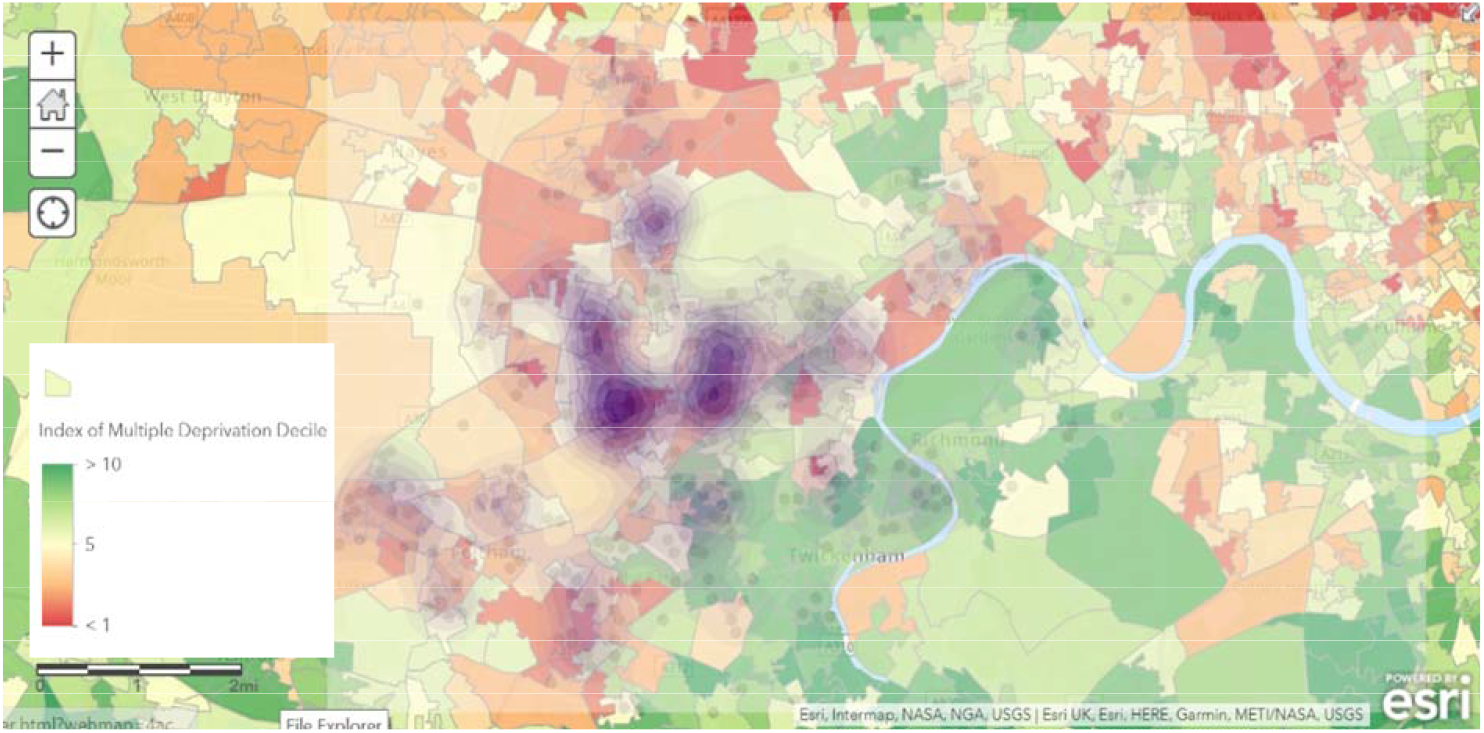
Covid-19 admission density and Indices of Multiple Deprivation as per 2015 data.

Household spread is postulated to be a significant factor in the onward transmission of disease. Case density appears to be greater in areas where the average household size is more than 2.1 people per household (Figure 5). There are limited areas with less than 2.1 persons per household within the catchment area but these appear to demonstrate relative sparing. It is not possible from this analysis to tell whether increased case density is a surrogate for increased population density in these areas there is an association with those thought to be in ‘poorer health’ and requiring admission. When figures 4, 5, and 6 are compared, areas can be identified with a lower density of cases corresponding to where overall household sizes are larger but deprivation and poor health are lower. We suggest that general health status and deprivation may be a greater indication of Covid-19 transmission than household size.

**Figure 5:**
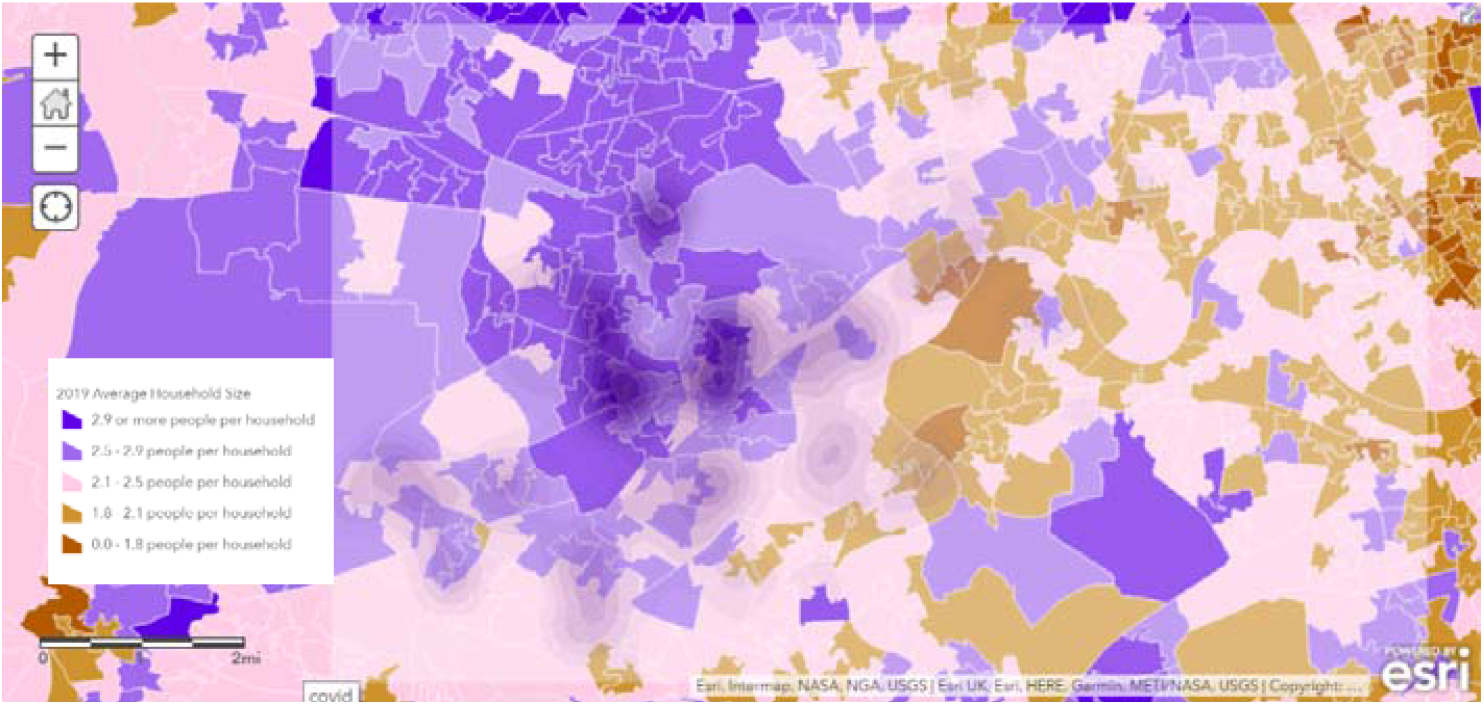
Household size and COVID-19 admission density.

**Figure 6:**
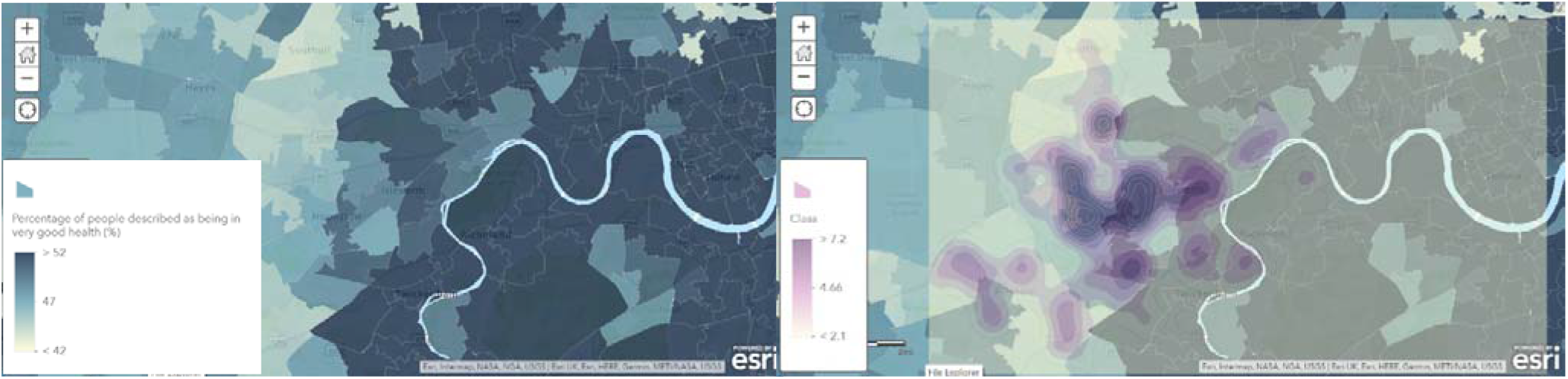
Self perceived health as per the 2011 consensus and COVID admission density.

## Discussion

This study demonstrates clusters of cases of Covid-19 requiring admission within the local hospital catchment area. This shows there is a role for targeted public health measures in tackling the spread of coronavirus in the community, and we go on to discuss these in more detail below.

### Areas of Deprivation

There is a well established link between health and social inequalities, examined by the Marmot report released in 2010[4]. In the decade since the original report there have been little improvements made[5]. Our study adds to increasing evidence, prevalence of Covid-19 is higher in those from more deprived areas of society [6]. This raises the question of whether severe Covid-19 is acting as an indicator of ongoing regional health inequality or whether these populations are at increased risk. There are several possible explanations for this; increased population density propagating spread of infection; poorer overall health making individuals more susceptible or presenting later to hospital[7]. Those from low income households have been found to be less likely to be able to work from home which may influence self-isolation[8].

Indices of deprivation are derived from 7 factors – income, employment, education, health, crime, barriers to housing and services, and living (a measure of the quality of indoor and outdoor environment. The UK is divided into Lower-Iayer Super Output Areas (LSOA’s) and are ranked from most deprived to least deprived. Lowest decile ranked LSOAs are the most deprived areas. In the areas covered in this study, Hounslow has 16 LSOAs in the top 2 deciles. All of the LSOAs in Ealing are ranked in the top 50% most deprived areas, whereas 90% of the LSOAs in Richmond are ranked in the 50% least deprived. This corresponds with our finding of increased density of cases in in Hounslow, Isleworth, Feltham and Southall (town centres in the boroughs of Hounslow and Ealing) with relative sparing in Richmond [3,9]. Baena-Diez et al showed an inverse relationship between mean household income and incidence of Covid-19 in Barcelona which appears to support our findings[10]. Similarly in France, an associated between higher deprivation and mortality was also seen [11]. Contrary to initial claims Covid-19 was a “great equaliser”, further UK research found disadvantaged communities are disproportionally affected highlighting health inequalities across socioeconomic groups[12].

### Household size

Viral spread within households has been highlighted as a prominent mechanism of transmission[13,14]. This has led to advice in the UK to limit indoor mixing to 6 people with a maximum of 2 households. Higher density of cases are seen in areas of higher household occupancy within the area studied, however, the upper limit in the mapping is >2.9 persons per household. It is possible there are areas with average household size significantly greater than this. This likely excludes multigenerational households where there may be a greater risk of hospitalisation due to older relatives within in the home[15] and younger household members who may be less likely to socially distance compared to the older age groups[16].

### Overall health

Figure 6 demonstrates there is a correlation with lower overall health and hospital admissions with Covid-19. A number of comorbidities have been implicated as risk factors for more severe illness [17], in particular cardiovascular disease and diabetes. Not only this, but it is reasonable to expect that patient’s with poorer overall health may have a lower physiological reserve to be able to combat an acute illness such as Covid-19. Patients with poorer overall health may also be more likely to come into contact with healthcare services increasing their risk of nosocomial infection[18].When interpretation of these maps is combined, areas can be seen with >2 persons per household that do not have increased case density. These areas appear to correlate with areas of less deprivation and greater overall health suggesting that these may have a greater influence on Covid-19 admissions.

The spread of data suggests that in the event of a second wave there may be a role for more intensive public health intervention and support in areas of greater deprivation and greater age. It may me argued that hospitals in areas exhibiting greater deprivation, poorer overall health and older age populations may be expected to be under the most pressure from Covid-19 admissions. This is in addition to the greater overall healthcare needs of these populations.

Our initial aim was to highlight areas for targeted public health intervention. UK-wide lockdown was introduced on 23rd March. The data shows significant reductions in admissions per day following this, demonstrating the success of widespread Public Health measures. However, the implementation of lockdown measures has created an inevitable economic downturn which will further increase the disparity in indices of deprivation across the UK. This will only further disadvantage those populations that are most at risk of Covid-19 [19].

There were high rates of Covid-19 admissions seen throughout hospitals in the first wave of the pandemic. The largest study to date suggests only up to 6% of the population have contracted coronavirus. Moreover, the effect on immunity is yet to be seen with possible reports of re-infection increasing [20,21].

### Limitations

The study demonstrated clusters of cases in Hounslow, Isleworth, Feltham and towards Southall with relative sparing in Richmond. This could otherwise be explained by patients from this area choosing to attend another hospital or ambulances diverting patients to other nearby hospitals which may be influenced by travel time and/or waiting times. By the same token, hospitals north of ours were under particular strain at the time of the study resulting in patients seen from outside the hospital catchment area. Maps drawn have been limited to the immediate geographical area of the hospital. Outlier cases also occurred from the south coast to the Midlands and were not included to allow a more detailed analysis of the local area. These presentations may be explained by the proximity of the hospital to a major international airport.

This is a single centre study and there it’s external validity is limited. This study solely focuses on patients with moderate to severe Covid-19 requiring admission to hospital. It was not possible to collect data regarding asymptomatic or mildly symptomatic patients due to a policy of only testing those requiring admission due to national testing strategies at the time of the study.

**What this paper adds**

- **Demonstrates hotspots of cases of coronavirus within the community**.
- **These are associated with areas of deprivation**.
- **Adds to further evidence there is a role for targeted public health measures especially those in most deprived areas**.

## Data Availability

Raw data is available from author.

## ACKNOWLEDGEMENTS

Maps generated using ArcGIS by esri.

---

Figure 4 living map layer provided by Esri UK based on data 2015 Data from the Department for Communities and local Government.

Figure 5 living map layer by <u>Michael Bauer Research</u> provided by Esri

Figure 6 living map payer procided by Esri UK based on information from the 2011 census, provided by the office for National Statistics

